# Perception and knowledge of artificial intelligence in healthcare, therapy and diagnostics: A population-representative survey

**DOI:** 10.1101/2022.12.01.22282960

**Authors:** Cornelius G. Wittal, Doerte Hammer, Farina Klein, Joachim Rittchen

## Abstract

Artificial intelligence (AI) is understood as a system’s ability to correctly interpret and learn from data, and to achieve specific goals and tasks through flexible adaptation to those learnings. Despite a broad range of available applications for artificial intelligence in medicine, healthcare professionals are reluctant to implement AI-powered devices. Data on the perception of medical AI in the German general public are currently rare. Therefore, two online surveys were conducted in 2021 in Germany to assess knowledge and perception of artificial intelligence in general and in medicine, including the handling of data in medicine. A total of 1,001 and 1,000 adults, respectively, participated in the surveys. The survey results stress the need to improve education and perception of medical AI applications by increasing awareness, highlighting the potentials, and ensuring compliance with guidelines and regulations to handle data protection. This survey provides first insights into this relevant topic within the German population.

## Introduction

First described by Alan Turing in 1950 as similar to but more complex than the human brain [1], artificial intelligence (AI) is nowadays understood as a system’s ability to correctly interpret and learn from data, and to achieve specific goals and tasks through flexible adaptation to those learnings [2]. The earliest work on AI in medicine dates back to the early 1970s [3]. Seminal advancements have been made since then, leading to today’s broad range of available applications for artificial intelligence in medicine [4]. These include smartphone-related monitoring systems, e.g. subcutaneous glucose or electrocardiogram (ECG) monitoring [5,6] and risk prediction of disease progression and development, e.g. prediction of glomerular filtration rate (GFR) decline in kidney disease or prediction of outcomes in gastrointestinal bleeding [7,8]. Furthermore, AI has been implemented in supporting cancer diagnosis using computational histopathology [9] and facilitating treatment decisions to improve patient care [10,11] and drug discovery research [12].

Despite promising areas of application, healthcare professionals, in particular physicians, show a certain reluctance to implement AI-powered devices due to various reasons including unpreparedness, administrative burdens, or lack of a legal framework or privacy issues associated with the use of AI [13,14]. According to surveys among Korean doctors and German undergraduate medical students, respondents considered the potential of AI to be limited in unexpected situations and believed that AI will not replace their roles in the future [15,16]. In addition to the attitude of healthcare professionals, the public’s perception of medical AI is crucial, as it may impact the development of AI products in terms of collecting sufficiently big data sets from the public for machine learning for instance. Content analysis of social media data in China revealed that negative public attitudes toward medical AI are often based on lack of trust in AI and the absence of the humanistic care factor [17]. However, data on the perception of medical AI in the German general public are currently rare.

Therefore, two online surveys were conducted in Germany in order to gain information on public awareness and knowledge, perception, fears and hopes towards AI in general and medical applications in particular. In addition, people’s perception of data generation and use in a medical context, e.g. willingness to share data, was investigated.

## Material and Methods

Two online surveys were conducted in November 2021 with identical procedures but different samples. Both surveys were unrelated to drugs or clinical trials and focus on the public perception and knowledge of either Artificial Intelligence or data usage in healthcare, therapy, and diagnosis (please see supplement for complete questionnaires). Participants from Germany aged >18 years anonymously completed online questionnaires focussing on artificial intelligence in medicine and use of data in medicine. Participants were recruited under the use of the Access Panel of the market research service provider Dynata. Samples for both studies were collected using nationally representative quotas for age, gender, and federal state. Soft quotas were collected on educational attainment. The surveys were performed in a classical, completely anonymous market research setting. After providing information about the purpose of the surveys the respondents were asked for their willingness to participate. Each participant received a unique ID. Duplicate participation was excluded by checking whether an ID occurred more than once in the data set. Participants either answered questions as free text, or ranked their perception on 7-point Likert scales ranging from -3 (strong rejection) to 0-1 (neutral) to 3 (strong endorsement), or their attitude from 1 (fully disagree/no interest/very bad) to 7 (fully agree, highly interested/excellent). In addition, sociodemographic aspects such as gender, age, type of school qualification, marital status, number of children, place of residence (German federal state), population of place of residence, type of residential area, and number of people in the same household were assessed.

Data from the surveys were analysed descriptively. Incomplete questionnaires/data sets were not included in the data set or in the analysis. For continuous variables, statistic parameters including arithmetic mean and range were calculated. Frequency distributions for discrete variables were provided as percentage in relation to the total sample. Free text answers were transferred post hoc into adequate coding schemes and analysed as frequency distribution. The survey assessing use of data in medicine was also conducted in 2020. Responses from the present survey were compared to those obtained a year earlier.

## Results

### Knowledge and perception of artificial intelligence in general

A total of 1.001 adults (51% female) participated in the survey about AI in medicine (Tab. 1). Asked to define artificial intelligence and its potential applications, 23% of participants provided a clear definition of AI as self-learning, optimising, autonomously acting systems. Top unprompted answers also included general support or takeover of work, computer-/ algorithm-based processes, and robotics (16% each). One out of 5 participants indicated no level of knowledge or didn’t provide any information.

**Table 1:**
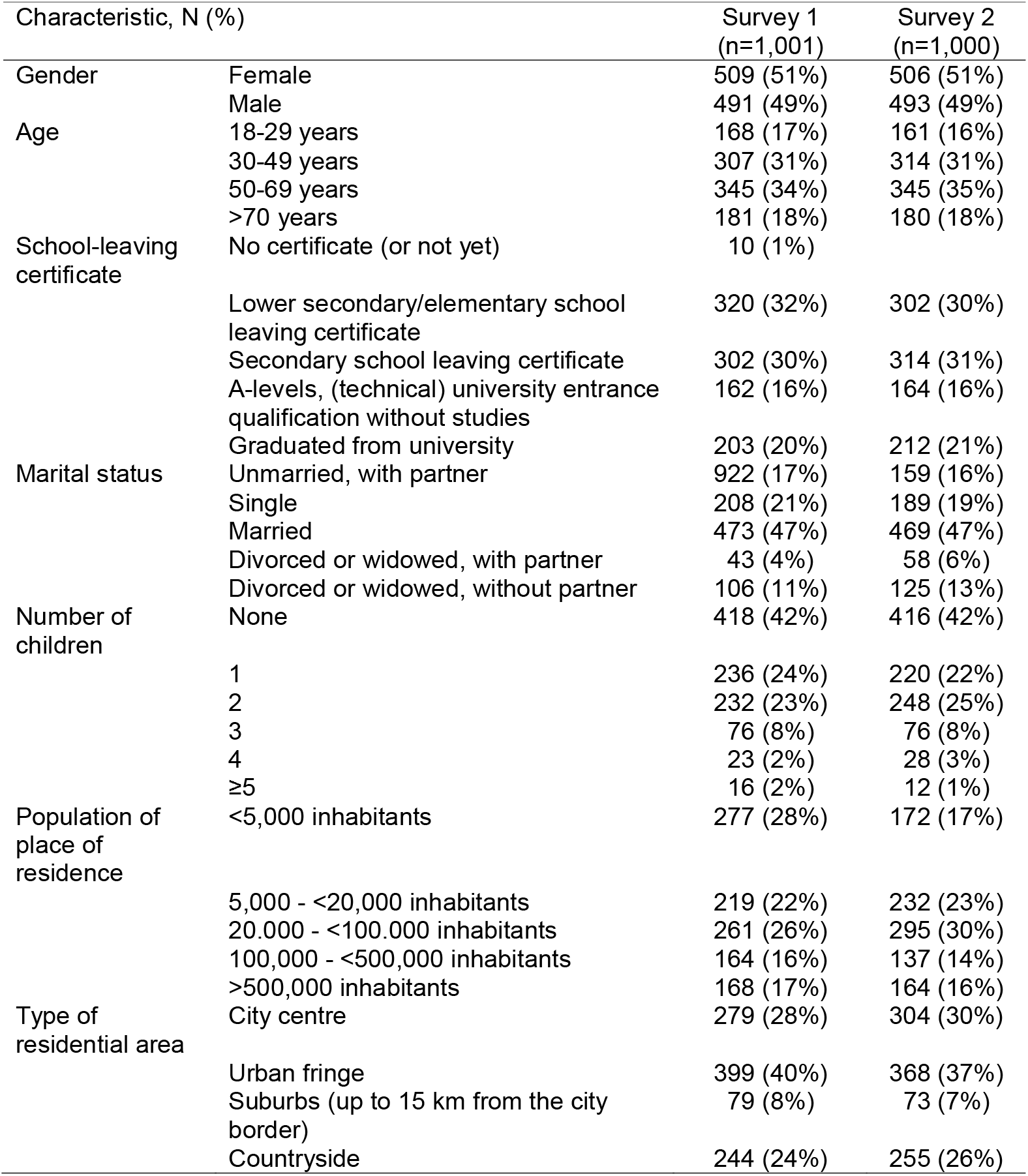
Demographic characteristics.

In addition to the level of awareness, perception of AI was also queried (Fig. 1A). Rankings revealed that 59% of participants had a neutral attitude towards AI in general, while 24% approached the topic positively. A negative attitude to the term AI was present in 17%. Taking a closer look at the perception of AI by ranking various feelings and attitudes, many participants felt interested (59%), curious (55%) and optimistic (50%), while 17-38% mentioned negative feelings (e.g. no interest, pessimistic) (Fig. 1B). In particular, fear (27%) and distrust (38%) were associated with AI. Less than half of the participants (45%) felt supported/supervised. In general, AI was perceived more positively by the group of participants with a high level of educational background compared to lowly educated participants.

**Figure 1:**
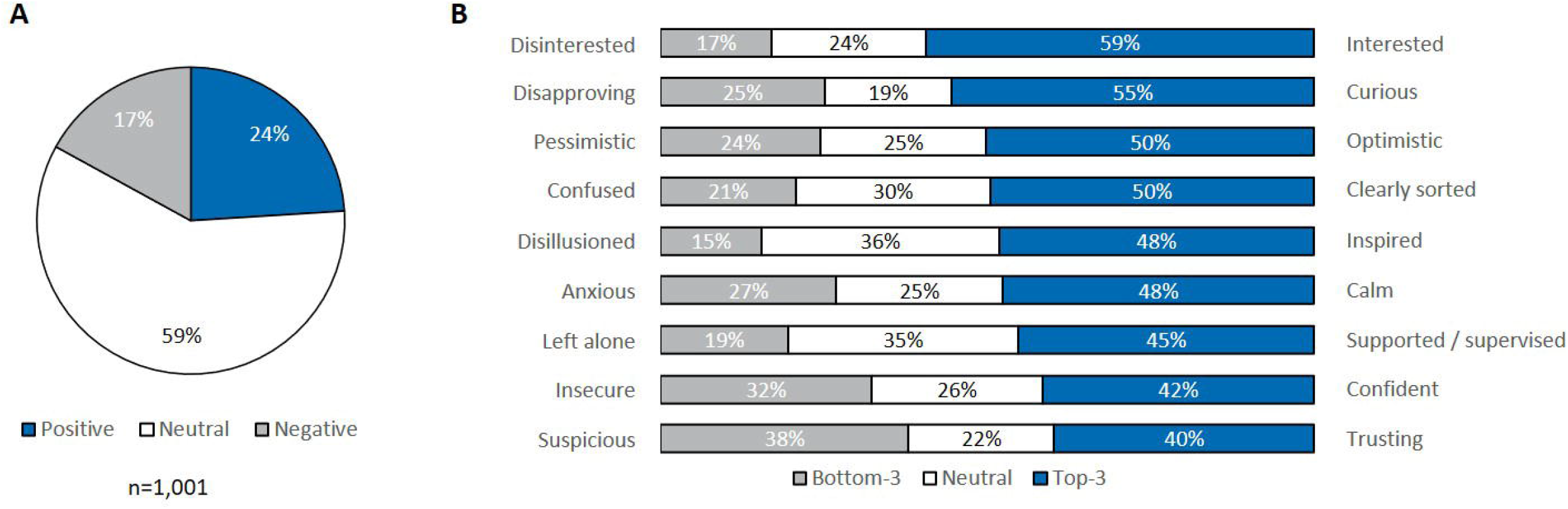
Perception of AI. Basic attitude towards artificial intelligence (A) and rating of feelings/attitudes (B). Feelings and attitudes towards AI were rated on a 7-point Likert scale; bottom-3, ratings -3 to -1; neutral, 0 to 1; top-3, 2 to 3

Participants were also specifically asked about their personal perception (agreement/disagreement) of suggested positive and negative connotations of special issues regarding AI. They often agreed upon positive connotation, for instance, AI may help to strengthen international competition and productivity of companies (67%), to make data-based decisions (63%), to explore complex relationships not manageable by humans (57%) or to assist with/execution of routine work (56%) (Fig. 2). In this setting, particularly participants with a positive attitude towards AI (n=243) agreed with positive connotations. At the same time, suggested negative connotations applied to many participants, with 59% agreeing that AI will replace employees and 71% felt that AI could have the disadvantage of dependence on programming specialists (Fig. 3). Those who predominantly rejected AI (n=171) were particularly afraid of replacement of human work by AI (78%), while this risk is also seen by 56% of participants with a positive attitude towards AI.

**Figure 2:**
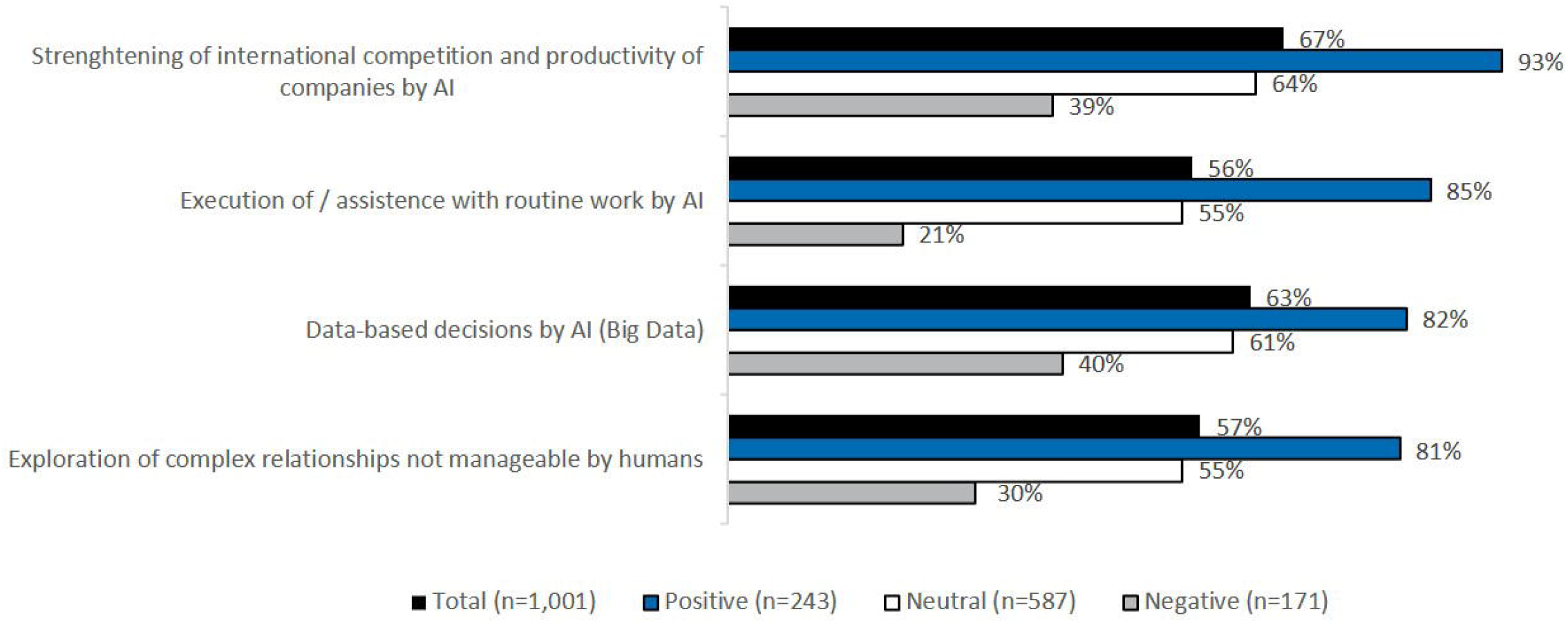
Agreement on potentially positive connotations depending on perception of AI.

**Figure 3:**
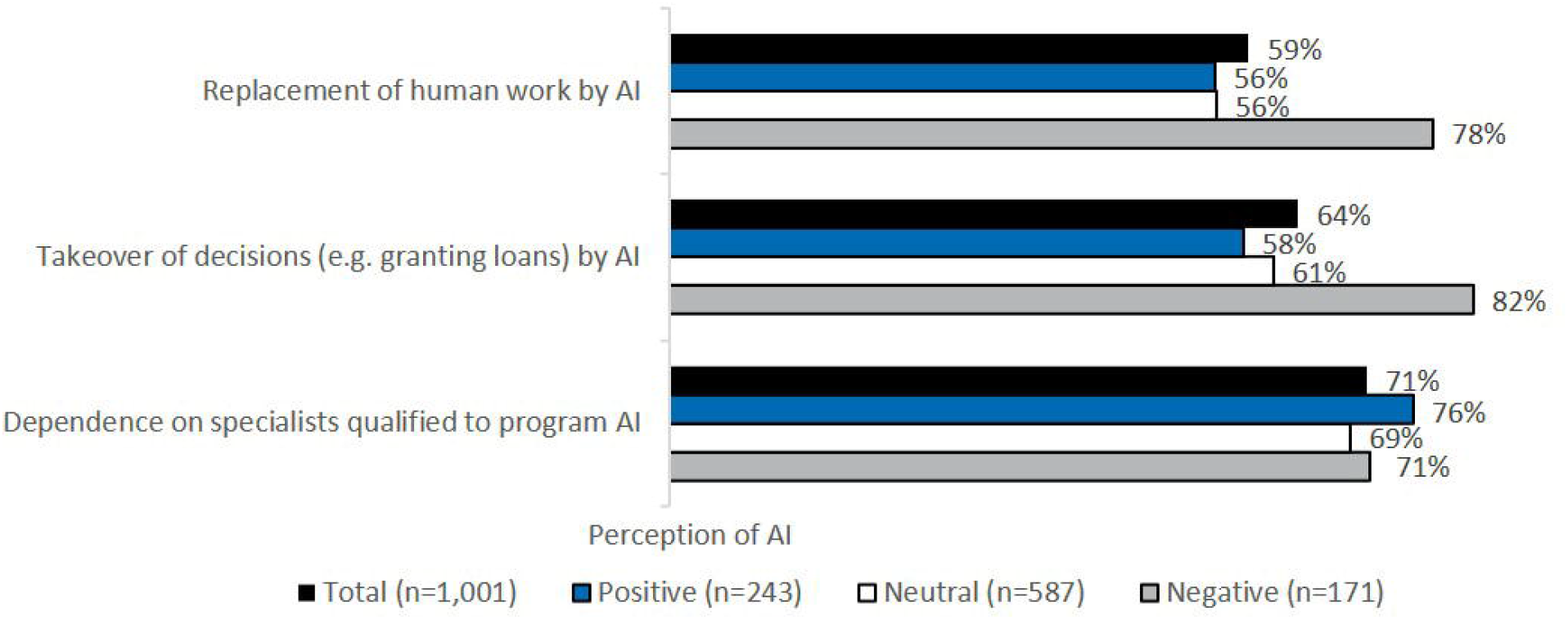
Agreement on potentially negative connotations depending on perception of AI.

### Knowledge and perception of artificial intelligence in medicine

While only 6% of the respondents gave unprompted examples of medical applications relying on AI, 56% of participants have encountered AI in the context of medicine and/or medical research when specifically asked about it. Among these (n=561), participants most frequently unprompted listed assistance during operations (40%) and imaging diagnostics (21%) as potential medical areas of AI implementation. Fewer participants mentioned data collection and evaluation (7%), new treatment methods (6%) or medical research and drug discovery (6%). However, more than half of the participants stated that they knew various predefined fields of implementation only by hearsay, while a maximum of 20% specified to have detailed knowledge, mostly independent of educational background (Fig. 4). The largest proportion of participants with detailed knowledge was found in the group of participants who had a positive perception of AI in general.

**Figure 4:**
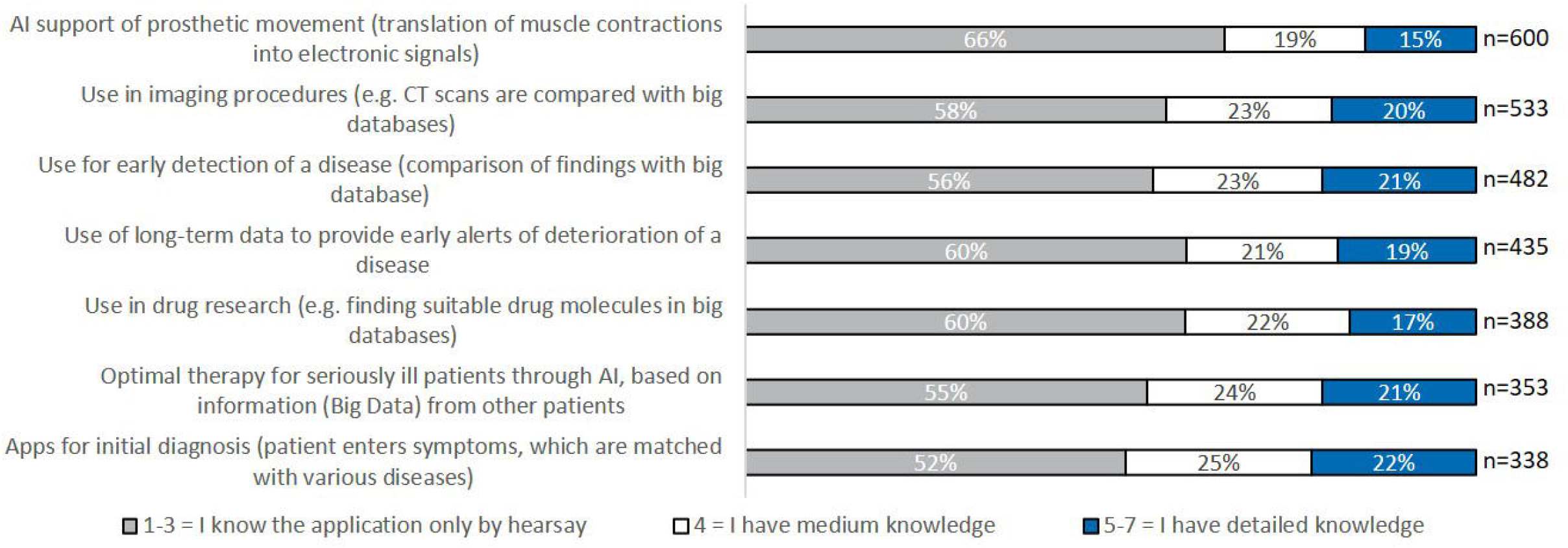
Knowledge on various predefined implementations of AI. Participants knowing the specified application filed rated their knowledge in pre-specified areas on a 7-point Likert scale (1-3, I know the application only by hearsay; 4, I have medium knowledge; 5-7, I have detailed knowledge)

The majority of participants supported the statement, that the use of AI in medicine could be “a real blessing” for diagnosis and therapy (72%) and that data protection should be revised with strict monitoring to prevent misuse, while enabling big data acquisition (70%). More than 3/4 of participants agreed that more information should be provided about possible applications of AI and that far too little is currently known. The readiness to share their anonymised health data to help building databases that AI can then use to improve diagnoses and therapies for seriously ill patients was expressed by 69% of participants. The percentage of participants agreeing with the aspects mentioned above increased to up to 94% when selectively considering the group of AI proponents.

More than half of the participants stated that they want to learn more about AI in medicine (59%), while 19% showed minimal or no interest (Fig. 5A). Among respondents who indicated moderate to high interest in the topic of AI (n=811), documentaries on TV and articles with pictures and interviews were considered as most suitable channels for generating awareness of the topic of AI in medicine (Fig. 5B). Online trainings, websites of organisations, audio podcasts or internet blogs were rated as a suitable educational option by 35-43%.

**Figure 5:**
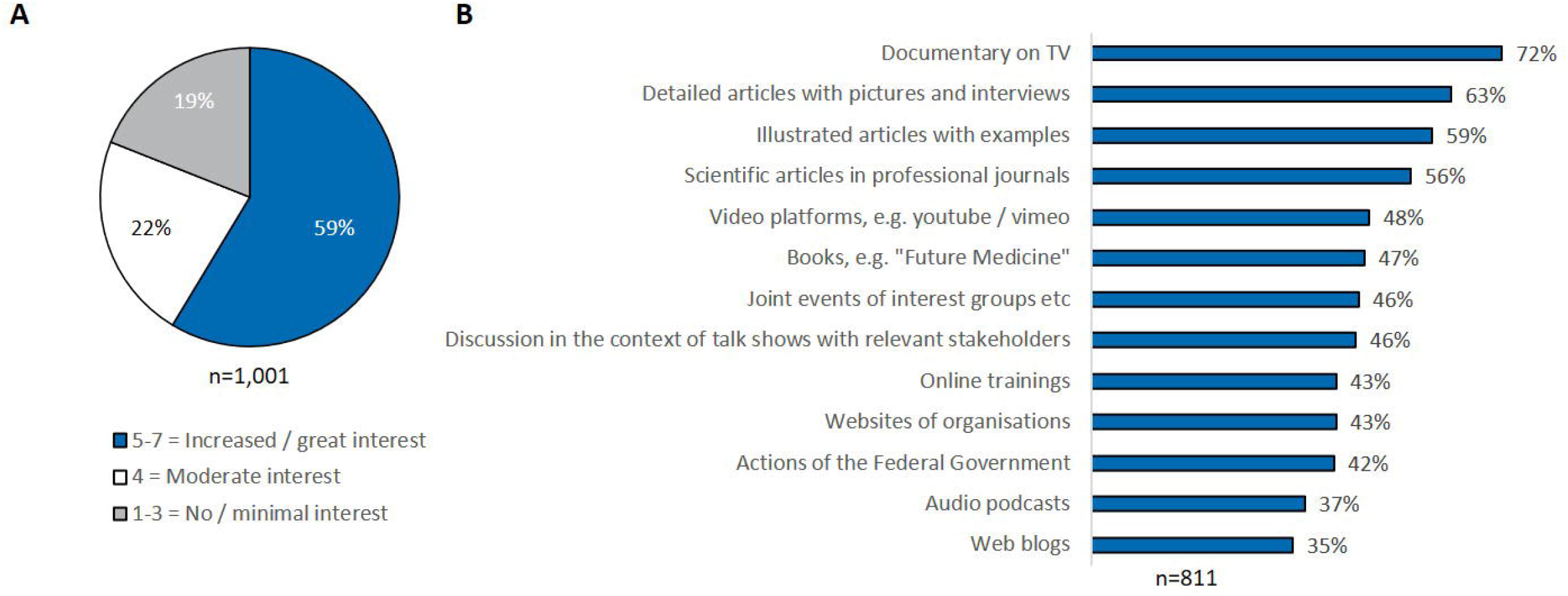
**A. Level of interest in learning more about AI** (all participants; 1-3, no/minimal interest; 4, moderate interest; 5-7, increased/great interest); **B. Preferred channels of education** (group of participants with medium to high interest)

### Perception of handling data in medicine

A total of 1,000 people participated in the second online survey about use of data in medicine. This second survey on the knowledge and perception of data relevant aspects in medicine revealed that 70% of the participants think about or tend to think about data privacy in their daily life (Fig. 6A). Compared to the 2020 survey, more participants care about data protection. Almost 40% of the participants had limited or no knowledge of where and with whom their personal data is shared (Fig. 6B). The level of knowledge about data protection was very low for 15%, very good for 35%, and medium for 50%. The top 3 unprompted responses to the question where big data sets are applied or generated in a medical context entailed consultations (36%), contact to health insurance companies (27%) and hospital stay (16%).

**Figure 6:**
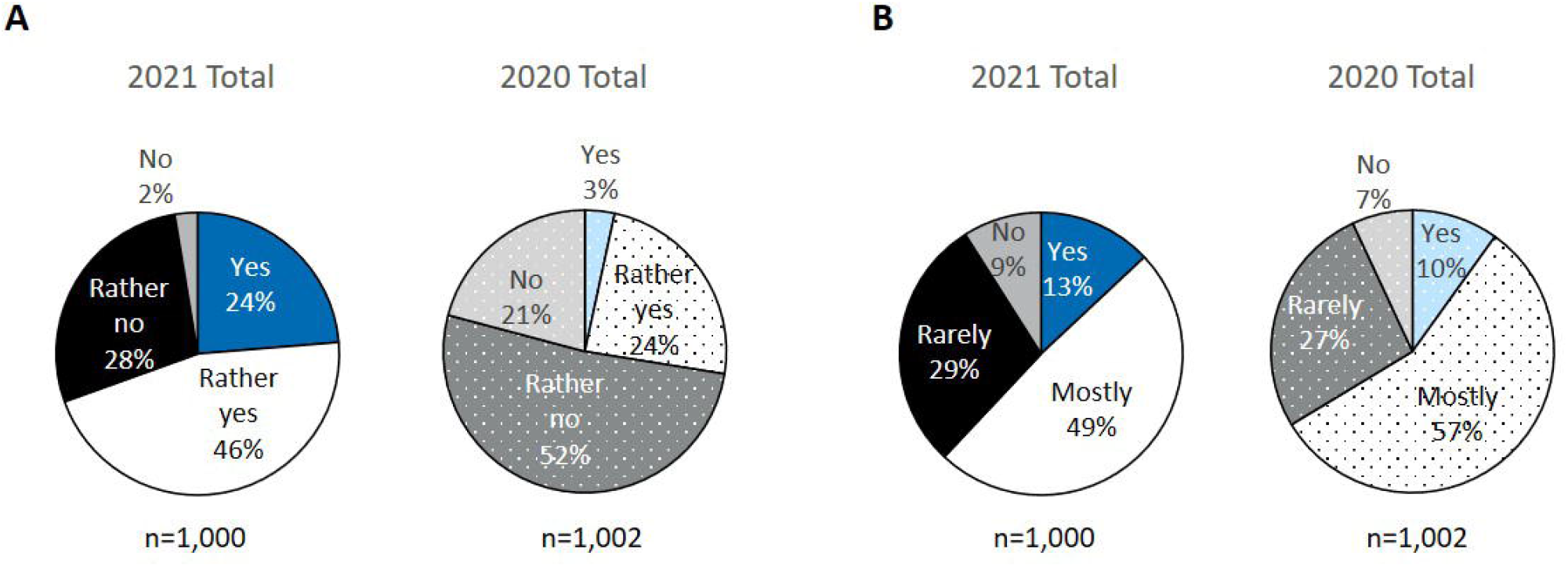
Data protection 2021 survey versus 2020 survey. A. Proportion of participants thinking about data privacy in daily life. B. Awareness of whom data is shared with

Developments such as data storage for better classification of current illnesses, health cards with personal access to clinical findings or the use of patient data for further development of therapies were perceived as most beneficial (Fig. 7). Half of the participants saw an advantage in telemedicine and online consultations, with a rising proportion to 71% in the group of people who had a positive attitude towards digitalisation.

**Figure 7:**
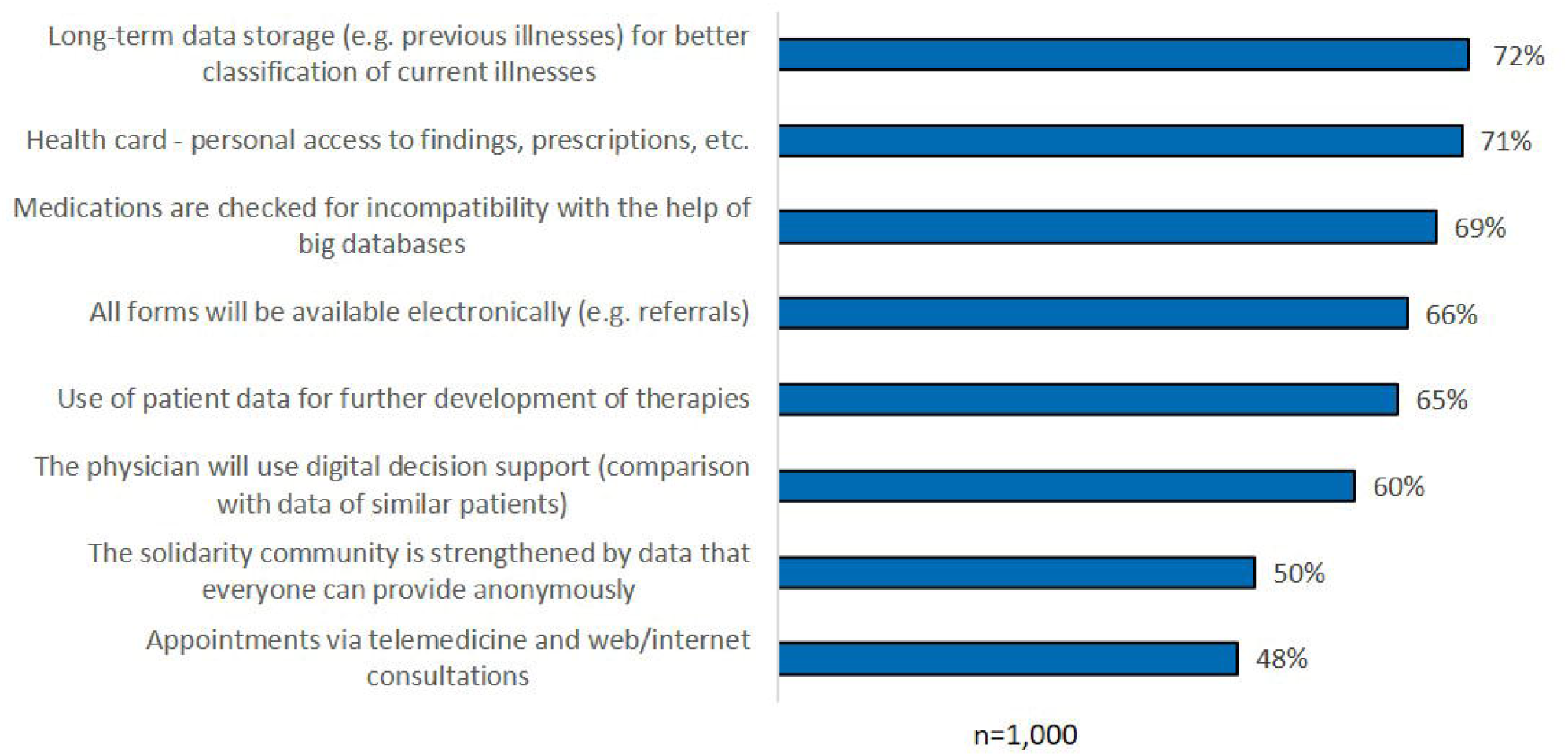
Perception of AI implementation as an advantage.

Participants were also queried which of the mentioned data-based applications in the health sector they were familiar with. Three out of 4 participants (77%) were aware of apps aimed at preventing the rapid spread of infectious diseases such as COVID-19, and 61% of apps for medication or exercise. Compared to the previous year, the level of awareness of COVID-19 apps increased (plus 4 percentage points), as did the level of awareness of digital pathology, i.e. disease findings from tissue samples through comparisons with large databases (plus 6 percentage points). Considering the group of digitalisation proponents, knowledge of potential data applications in health care is higher compared to people who have a negative attitude towards digitalisation.

Preferred addressees of data for new data-based applications in healthcare were also examined (Fig. 8). The willingness to pass on data to public institutions such as health insurance companies (73%), health authorities (57%) or universities (50%) was higher than the willingness to entrust data to private companies. Here, 36%, 25% and 22% would pass on their health data to medical technology, pharmaceutical and IT companies, respectively. Overall, the willingness to share data has increased compared to the previous year. The younger the respondents, the higher was the willingness to share data. Likewise, the group of participants with a positive attitude towards digitalisation showed a higher willingness to share data. Within the group of patients who would share their data with the pharmaceutical industry, there was generally a greater willingness to pass on data also to other addressees. Distrust (33%), profit orientation (18%) and lack of data protection (14%) were mentioned unprompted as reasons for not sharing data with pharmaceutical companies. The majority of participants (63%) indicated that they would be willing to provide a pharmaceutical company with their anonymised treatment data for further research if they responded to the company’s drug in the event of a serious illness.

**Figure 8:**
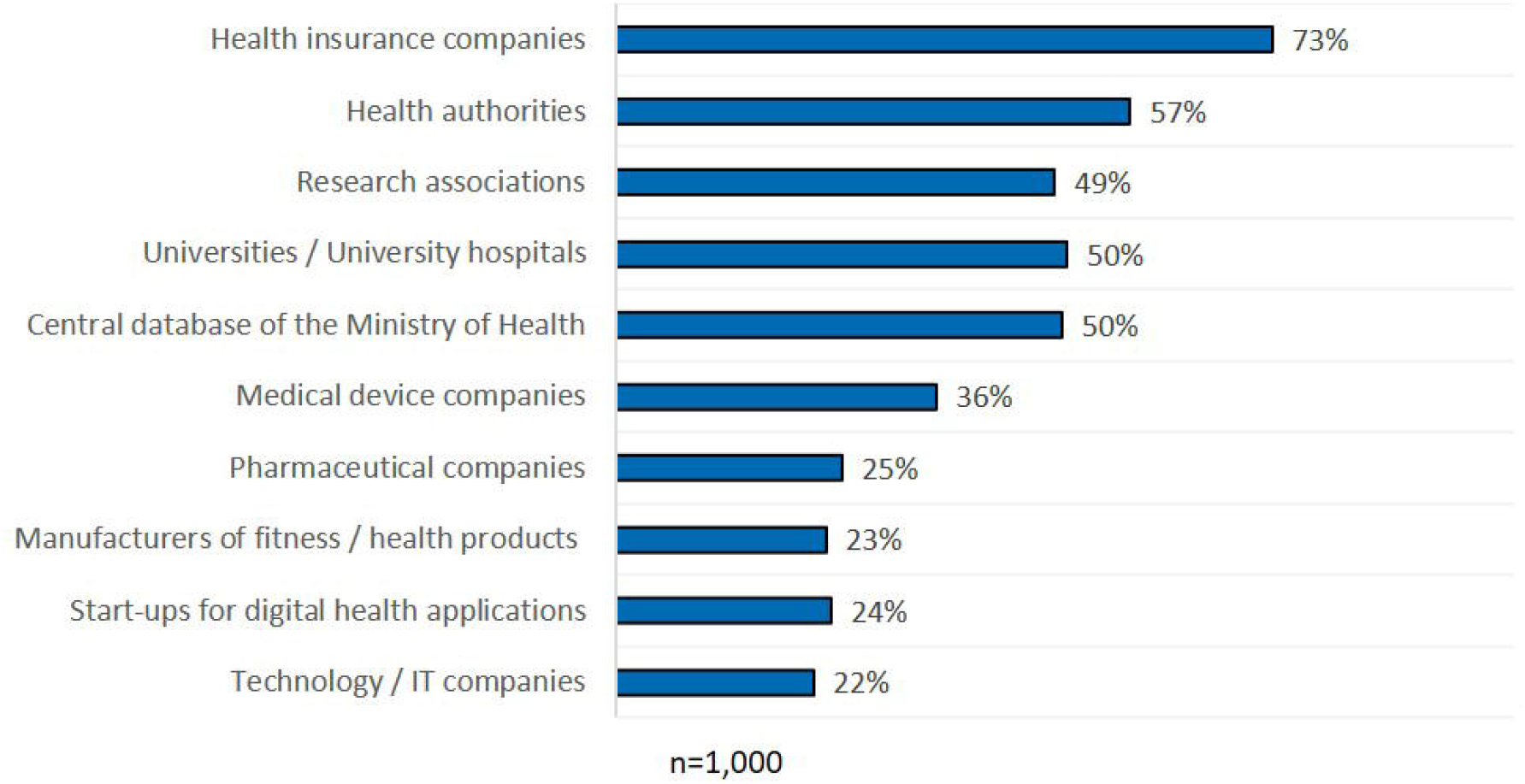
Readiness to share data with pre-specified addressees.

## Discussion

Since the first conceptual idea of machine learning and artificial intelligence in the middle of the last century, great progress has been made in many areas. Within medicine, artificial intelligence-powered technologies are already applied and rapidly evolving [13]. But despite this, most people do not recognise the technological efforts and how they actually affect everyone’s life. The current surveys on the knowledge and perception of artificial intelligence and data generation and use in medicine included participants representative of the German population based on distribution of gender, age and educational background as well as place of residence [18-20]. They demonstrate the general German public’s low level of knowledge, as only 6% of the respondents unprompted mentioned medical approaches when asked for AI application areas. However, a higher educational background of the participants correlated with increased knowledge of medical AI. Similarly, a survey conducted in 2021 reported a low level of digital health competence in the German general population, in particular in the subgroups of senior citizens and people with lower educational background [21]. As well, the general perception of AI was associated with the level of knowledge. In the subgroup of respondents supporting AI, twice as many people indicated high levels of knowledge compared to the ones with neutral or even negative attitudes towards AI. Nevertheless, it is likely that the majority of the 1,000 respondents already had contact to AI-supported medical approaches, such as robotic-supported surgeries, radiological examinations or newly designed drugs, since AI is already integrated in different medical fields as well as drug discovery [22].

Regarding the recent advances, medical AI is likely to help streamline diagnosis and disease prediction, reduce medical errors, and improve decision-making and therapy [17,23]. In order to exploit the full potential of emerging AI products, the public’s perception of medical AI is crucial as it would require collection of big data sets from the public [17]. While people with a higher educational background are more likely to specifically search for more detailed information and consequently gain a more positive perception of the topic, it is harder to reach and convince people with middle or low educational background. Yet, in line with findings of a Chinese content analysis of social media data [17], the survey revealed a high interest in learning more about medical AI by the majority of participants independent of their educational background.

Hence, there is opportunity to increase the perception of the public for AI in medicine and health care by different educational approaches. One of these approaches are cooperative information projects between public journals and pharmaceutical companies targeting the interested people. Additionally, there are few online platforms offering information and courses on AI in general and AI in medicine particularly, such as the AI Campus and the Platform for Learning Systems [24,25]. Although these platforms do not just address the higher educated people, the majority of the population might not be reached. To find the platforms, time-intensive research is needed requiring the intention to learn more about AI. The importance to use appropriate channels for education was underlined by the survey displaying easy-to-understand formats in television and magazines as the leading media of choice for generating awareness. Specialist events or publications are less popular in the public based on the respondents’ answers. Nevertheless, they should still be maintained as educational source because previous studies revealed also a need for better education of specialists, many of whom lack a full understanding of the principles of AI [14]. To ensure the correct application of AI requires trained healthcare specialists, such as physicians, surgeons, but also caregivers [26], however the existing training offers are few and not easy to find. For example, in May and June 2021 only 30 out of 87,136 CME-certified courses featured AI-related titles [27]. There is a strong need for more courses focusing on AI, which should be also CME-certified. In this context, two courses from the online platform AI Campus were recently officially recognised for medical training following cooperation with the Baden-Württemberg State Medical Association.

While the majority of health professionals recognised AI as useful in their field, still many physicians exhibit clear reservations towards AI, including serious privacy issues [13,14]. There is a lot of controversy on the subject of AI, not only within the scientific and medical community, but also among the public, being divided on potential benefits and risks of AI [14]. This was also reflected in the survey, since a high percentage of participants agreed on both positive and negative connotations associated with AI, simultaneously displaying fear and hope. Similar to the perception of the specialists, the respondent’s fears mainly are based on the important aspect of data protection and privacy issues.

Health privacy poses major legal and ethical challenges when dealing with big data sets [28]. Within the EU, the General Data Protection Regulation (GDPR) harmonised data protection and privacy laws across the member states and enhanced data security and privacy regulations. Security when processing personal data in medical research is regulated by the options for anonymisation and pseudonymisation. While pseudonymisation describes the replacement of personal data with a code or number, for example, anonymisation refers to deletion of personal data. Hence, pseudonymisation means that the person responsible for processing personal data cannot identify the person, but by adding information it would be possible for third parties to disclose the identity. By anonymising personal data, this re-identification is not possible, neither by the responsible person nor by any other third party [29]. Although pharmaceutical companies in general do not have access to personal patient data, the survey showed more scepticism towards sharing data with the industry than with government institutions. A Chinese content analysis of social media data came to similar findings, showing that some people had negative attitudes toward medical AI due to a general distrust of AI companies [17]. A market research survey conducted in Germany indicated that 80% of respondents did not oppose making digitally collected health data, for example with a fitness bracelet, their smartphone or other devices, accessible for medical research [30]. To enable better use of data for research and thus progress in prevention, diagnostics and therapy, the German Society for Internal Medicine e.V. (DGIM) calls for the adjustment of data protection regulations in Germany [31]. Once again, the key to a better public perception of disclosing data is education. The survey displayed a higher willingness of data disclosure for AI applications by respondents who showed higher interest in medical AI and supported digitalisation in general. In terms of direct self-benefit, participants demonstrated a higher perception of data sharing with pharmaceutical companies. Thus, the public should be informed about the potential personal benefits of medical AI through appropriate channels and easily understandable formats.

One example of beneficial medical AI is the cloud-based NAVIFY Tumour Board solution. The software platform facilitates the extraction of key data from clinical data and integrates relevant information into a single source. In addition to preparing information, the system also assists with presenting and documenting information. Hence, it is able to reduce the number of steps and time required for tumour board preparation and processing [32,33]. In addition to saving time and reducing costs, the platform also has the advantage of improving clinician decision-making through access to databases. The more data is disclosed in these databases and made available for use, the more precisely the AI-based systems can work.

As AI-mediated medical capabilities continue to advance, personalised medicine will also become achievable. Unique personal health data, such as genomic data could greatly advance the development of personalised medicine. While these kind of data always allows conclusions to be drawn about the identity of the donor, even if anonymised, the hurdle is to create very high data protection standards in order to ensure the positive public perception that is necessary for the collection of sufficient and meaningful amounts of data. Several projects and initiatives within the EU try to foster the use of genomic data to improve diagnostic or therapeutic options for patients, especially with rare diseases, such as “‘#WeWontRest” by the European Federation of Pharmaceutical Industries and Associations or “Screen4Care” by the Innovative Health Initiative [34,35].

The advantages of personalised medicine are rapid and accurate diagnoses as well as tailored therapies that lead to longer life with better quality of life can therefore potentially benefit the entire healthcare system. The path to personalised medicine will be closely linked to the digitalisation of medicine, which as well depends on the willingness of the public to share their personal data. Policy makers across Europe should increase their efforts to raise public awareness of privacy issues related to personal health data by involving them in data-driven medical AI development projects. One of these projects is the German initiative “genomDE”, which in association with the European genome initiative “1+ Million Genomes” aims to build up a platform for diagnostic genetic data for the improvement of research and health care [36,37].

Recently in Germany, another step towards a more digitalised medicine was made by the adoption of the “Act to Improve Healthcare Provision through Digitalisation and Innovation”, which included development and implementation of the electronic health card and electronic health record. Different personal and health-related data are stored on the eHealth card, such as diagnoses, prescriptions and therapy instructions, enabling that invoicing data available to the health insurance funds are collected in pseudonymised form at a research data centre, and for anonymised findings to be transmitted to research institutes upon request. This will make a larger amount of more current data available to science within a protected space, so that new findings can lead to improvements in healthcare. As health data is extremely sensitive, data protection provisions of the Fifth Book of the Social Code (SGB V) require adaptation to achieve optimal legal conditions for data protection [38]. To address prevailing public concerns, policymakers are called upon to develop a transparent and publicly comprehensible data security culture. In other countries, such as Denmark and the UK, people have more confidence in data protection. For instance, health data has been stored electronically for years in Denmark [39].

An obstacle to the digitalisation of medicine in Germany is the federal system with different regulations and laws between and even within the individual federal states. To address this issue, university hospitals in Baden-Wuerttemberg have joined forces [40]. Additionally, a consortium – the Medical Informatics Initiative – has been created across the borders of the federals states, aiming at improving the use of health data to ensure full benefit from digitalisation for every individual patient [41]. Efforts are being made not only in Germany and the EU to develop uniform and generally applicable data protection regulations with high security standards, but also worldwide. In 2013 the Global Alliance for Genomics and Health (GA4GH) was founded by genomic researchers, health care specialists, data scientists and other stakeholders to establish policy frameworks and technical standards for international sharing of health, genomic and other molecular data [42]. Despite aforementioned barriers, efforts by German policy makers to promote medical digitalisation can be successful. Thus, Germany is the first country in the world to prescribe digital apps (DiGAs) and reimburse them through the statutory health system [43]. In addition, the Federal Cancer Registry Act was amended so that from 2023 on clinical data from the cancer registries of the individual federal states will be combined in a nationwide registry [44].

However, policy makers, scientists, clinicians, but also patients and the general public still need to put a lot of effort into rapidly advancing the development of medical AI and, in general, the digitalisation of medicine. The current survey results stress the need to improve education and perception of medical AI applications by increasing awareness, highlighting the potentials, and ensuring compliance with guidelines and regulations to handle data protection. Ethical aspects should also be considered, in particular in highly complex and future-oriented fields, such as digital medicine [45]. In this way, people’s trust and acceptance regarding their willingness to share their pseudonymised health data with pharmaceutical companies can be gained. This might facilitate the execution of clinical trials, help to improve therapies, strengthen research and thus lead to achieving benefits for the entire society. In the future it is therefore advisable to not only consider the physician’s perception, but also to better understand the attitude of the public toward AI. This survey provides first insights into this relevant topic within the German population.

## Limitations

The reliability of survey data may depend on the motivation, honesty and encouragement of participants. Survey question answer options could potentially lead to unclear data because certain answer options may be interpreted differently by respondents.

## Supporting information

Supplement 1_Questionnaire data in medicine

Supplement 2_Questionnaire AI in medicine

Supplement 3_CROSS checklist

## Data Availability

The authors confirm that the data supporting the findings of this study are available within the article.

