## Supplement 1_Questionnaire data in medicine for "Perception and knowledge of artificial intelligence in healthcare, therapy and diagnostics: A population-representative survey"

**Questionnaire – Data in Medicine 2021**

| | |
|---|---|
| **Method:** | Survey via online access panel |
| **Sample size :** | n=1,000 |
| **Country:** | Germany |
| **Target group:** | Adults |
| **Quota:** | Nationally representative quotas for age, gender, and federal state, education (soft quota) |
| **Interview duration:** | Ø 15 min |

**Programming notes**

| | |
|---|---|
| **SP:* | *single pick* |
| **MP:* | *multiple pick* |
| **RD:* | *randomization* |
| **MA:* | *matrix* |
| **OQ:* | *open question* |
| **NQ:* | *numeric question* |
| **SC:* | *scale* |
| **RA:* | *ranking* |
| **DD:* | *drop down* |
| **EX:* | *exclusive* |
| *[SCREENOUT]* | *Not included in target group* |
| *[END OF INTERVIEW]* | *End of interview* |

**Welcome page**

Dear participants,

We are very pleased that you are participating in this survey on "data in medicine".

The survey will take approximately 15 minutes of your time. Participation is of course voluntary and your data will be evaluated anonymously, so that no conclusions can be drawn about your person. Data will be used for market research purposes only.

If you would like to participate in the survey, please click "Next" to start the survey.

### Sociodemographic & Screening

Q1) Please provide your gender. **SP*

1. Female
2. Male
3. Diverse / not specified

Q2) How old are you? **NQ*

_______ years *[Screenout <18]*

Q3) In which federal state do you live? **SP*

1. Baden-Württemberg
2. Bayern
3. Berlin
4. Brandenburg
5. Bremen
6. Hamburg
7. Hessen
8. Mecklenburg-Vorpommern
9. Niedersachsen
10. Nordrhein-Westfalen
11. Rheinland-Pfalz
12. Saarland
13. Sachsen
14. Sachsen-Anhalt
15. Schleswig-Holstein
16. Thüringen

Q4) What school qualifications do you have? **SP*

1. No certificate (or not yet)
2. Lower secondary/elementary school leaving certificate
3. Secondary school leaving certificate
4. A-levels, (technical) university entrance qualification without studies
5. Graduated from university

### Data sharing

Q5) Repeatedly the statement "data is the gold of the 21st century" is mentioned in interviews or read in magazines or newspapers. Or there is talk of an enormous wealth of data from which society can benefit.
If you think about your normal work, leisure and private life: Where do you share data (e.g. because you post something online)? *Please fill in everything you can think of and provide concrete examples if possible.* **OQ*

Q6) In your opinion, where or when data is used from other users when a service is offered to you? *Please fill in everything you can think of and provide concrete examples if possible.* *OQ

### Data and data use

There are a lot of opinions about data and the use of data. We have compiled some of them and would like to know to what extent you agree with the statements compiled.

Q7) The use of anonymous data leads to great service offers, e.g. when shopping online or when using search engines. *SC

1. Totally disagree
2. 
3. 
4. 
5. 
6. 
7. Totally agree

Q8) The internet displays the future and leaving data there is inevitable. That's why you should just keep up with the times and not worry about anything else. *SC

1. Totally disagree
2. 
3. 
4. 
5. 
6. 
7. Totally agree

Q9) I can mostly decide for myself which of my data is available online as information. And as many millions of users show: misuse hardly ever happens. *SC

1. Totally disagree
2. 
3. 
4. 
5. 
6. 
7. Totally agree

Q10) Online you have to be careful where you click and thereby quickly agree to the use of data. All of a sudden you get unwanted emails and suggestions that just annoy you. *SC

1. Totally disagree
2.
3.
4.
5.
6.
7. Totally agree

Q11) Fitness bracelets help me to maintain my health – I accept that the data is with the provider. ** SC*

1. Totally disagree
2.
3.
4.
5.
6.
7. Totally agree

Q12) There are smartphone apps that help to deal with an illness (e.g. taking medication or memory training exercises). As long only my treating physician can access my data, that's ok with me.**SC*

1. Totally disagree
2.
3.
4.
5.
6.
7. Totally agree

Q13) I know that I can influence the transfer of data in internet applications, e.g. I can prevent it. However, it is not always easy to find the appropriate settings within the application. **SC*

1. Totally disagree
2.
3.
4.
5.
6.
7. Totally agree

Q14) Do you generally have an overview of whom and where you share your data with? *SP

1. Yes, I always have an overview.
2. I usually have an overview.
3. I rarely have an overview of whom and where I am sharing data with.
4. No, I no longer have an overview of whom and where I share data with.

### Data protection

In the following, we would now like to learn a little about your opinion and experiences with regard to data protection.

Q15) A general question: Is data protection a topic that you think about a lot or that concerns you in everyday life? *SP

1. No
2. Rather no
3. Rather yes
4. Yes

Q16) How would you rate your knowledge on data protection? *SC

1. I'm not familiar with it at all
2. 
3. 
4. Partly
5. 
6. 
7. I know it very well

Q17) In your opinion, how is the topic of **data protection** handled in Germany? Is data protection insufficent, are the existing data protection laws sufficient or is there rather too much data protection? *SC

-3. Not enough data protection / too many gaps
-2.
-1.
0. Data protection is just fine
1.
2.
3. Too much data protection/hindering of activities

Q18) And how well informed do you feel about the subject of data protection? *SC

1. I feel very poorly informed
2. 
3. 

4.
5.
6.
7. I feel very well informed

The following questions and the rest of the survey deal with the topic of "data in medicine", i.e. the application and use of data on diseases and corresponding therapies in the healthcare system and the resulting benefit to society in general, but also to the individual patient.

Q19) Data is generated by everyone who has contact with the healthcare system, whether as a patient or as an insured person. If you think about the last few years regarding yourself or those around you and about such contacts with the **health care system** (e.g. health insurance company, doctor, hospital): Where do you think **data is used or even newly generated**? *Please fill in everything you can think of and provide concrete examples if possible. *OQ*

Q20) Listed below are various occasions in which you, as a patient and insured person in the health care system, use data, some of which is newly generated and processed. Which of these occasions do you already know or are aware of? *Please tick all the opportunities that you are aware of. *MP, RD*

1. Data as an insured person with the health insurance company (e.g. name, insurance number, membership rate, illnesses, costs incurred)
2. Prescription redemption in the pharmacy (e.g. insurance number, prescribed medicines, issuer of the prescription, place of redemption)
3. Medical records at the physician's office or in the hospital (e.g. name, insurance status, previous illnesses, previous and current drug therapies, therapies administered and treatment success in surgery or other non-pharmaceutical therapies)
4. Patient history with the health insurance company (e.g. name, insurance status, treatment history, place of therapy, treatment success)
5. None of the above **EX*

### Advantages: Digital health system of the future

Experts agree that the **healthcare system of the future** will be based on digital processes and will access data much more than today. However, there will also be **significant patient benefits** associated with this. We have listed some of these benefits below.
*Please tell us how you rate each of them: i.e. whether it is an advantage from your point of view or not.*

Q21) As a patient, I will have access to my treatment data such as findings and all prescriptions via my health card at any time and can decide by myself to which physicians I share my health data with. *SC

1. No advantage: I totally disapprove
2. 
3. 
4. 
5. 
6. 
7. Advantage: I totally approve

Q22) All forms will be available electronically (e.g. bank transfers) and can easily be sent to me by email or inserted into an electronic file (e.g. access via mobile phone app). *SC

1. No advantage: I totally disapprove
2. 
3. 
4. 
5. 
6. 
7. Advantage: I totally approve

Q23) My data is also available years later in order to draw conclusions from previous illnesses and to better classify possible associations with the current illness. *SC

1. No advantage: I totally disapprove
2. 
3. 
4. 
5. 
6. 
7. Advantage: I totally approve

Q24) The physician will use digital support in decision making. Comparing with data from similar patients can be used to decide for the best therapy for me. *SC

1. No advantage: I totally disapprove
2. 
3. 
4. 
5. 
6. 
7. Advantage: I totally approve

Q25) Telemedicine and doctor's appointments as personal web/internet consultations will become normal, so that I can often spare the time-consuming visit to the practice. *SC*

1. No advantage: I totally disapprove
2. 
3. 
4. 
5. 
6. 
7. Advantage: I totally approve

Q26) Since my medication data will all be saved, I can be certain that incompatibilities between drugs are excluded, as this is automatically checked in large databases with every new prescription. *SC*

1. No advantage: I totally disapprove
2. 
3. 
4. 
5. 
6. 
7. Advantage: I totally approve

Q27) The solidarity community will not only be reflected in the membership rates where people with high income help finance the rates of people with low income and families. It will also refer to data that I share anonymously so that the entire system can be improved. *SC*

1. No advantage: I totally disapprove
2. 
3. 
4. 
5. 
6. 
7. Advantage: I totally approve

Q28) The data obtained during treatment of other patients will be used beneficially for the further development of therapies and may benefit me in the event of my own illness. *SC*

1. No advantage: I totally disapprove
2. 
3. 
4. 
5. 

6. 
7. Advantage: I totally approve

### Use of data in healthcare

Q29) Below we have listed several **new applications of data in healthcare** beyond those listed in the previous questions. Technology companies and start-ups specialising in artificial intelligence (AI) and the analysis of large data sets (big data) are often involved. Which of the following applications are you familiar with? **MA, Row RD*

*Columns*

1. Is known
2. Is unknown

*Lines*

1. <u>Selection of an optimal therapy</u> for seriously ill patients using artificial intelligence (AI), based on treatment data and treatment course (big data) of other patients.
2. "<u>Digital Pathology</u>", i. e. Precise diagnosis based on your own tissue samples by comparison with large databases (big data) where the results of similar tissue samples from other patients are stored.
3. <u>Suggestions from the health insurance company</u> for more exercise or a change in diet based on your own medical history, any previous illnesses and other data stored by the health insurance company (e.g. age).
4. <u>Evaluation of treatment data from medical records</u> for various diseases in order to draw conclusions about the efficiency of diagnoses and subsequent therapies under real-world conditions (so-called healthcare research).
5. <u>Evaluation of your own movement behaviour</u> and comparisons with other data (benchmarks) from suppliers of <u>fitness bracelets/watches or from smartphone apps</u>.
6. <u>Smartphone apps</u> that help <u>prevent</u> the rapid spread of <u>infectious diseases</u> such as COVID-19.
7. <u>Health apps</u> that help you to take your medication on time or provide exercises that help you e.g. to train arms/legs after illness, eyesight or memory. Medication intake and results of exercises are transfered to the doctor's office.

*Filter: Show for all applications in Q31 (items in line) where in column=1 (is known) was elected*

Q30) Please indicate how familiar you are with this application/s using the scale below: **MA, SC, Row RD*

*Columns*

1. I only know the application from hearsay
2. 
3. 
4. I have an intermediate level of knowledge
5. 
6. 
7. I have detailed knowledge

*Lines: Applications similar to Q31, if known*

*Filter: Show for all applications in Q31 (items in line) where in column=1(is known) was elected*

Q31) As you have noticed in the previous questions, many applications are conceivable in which health data can be used. But use also requires that patients or insured persons agreement to pass on their data to third parties for evaluation and processing. For the applications listed as known in the previous questions: Would you be willing to share your data with third parties? **MA, Row RD*

*Columns*

1. Absolutely not
2. Maybe: It depends on who receives my data
3. Definitely yes

*Lines: Applications similar to Q31, if known*

Q32) Now in general: Which of the institutions/companies listed below would you or rather not entrust your own health data to? *MA, SC, Row RD

*Columns*

1. I would <u>definitely not</u> entrust data
2.
3.
4.
5.
6.
7. I would <u>definitely</u> entrust data

*Lines*

1. Health insurance companies
2. Health authorities (e.g. local health department)
3. Pharmaceutical companies
4. Medical technology companies
5. A central database of the Federal Ministry of Health
6. Digital health start-ups
7. Universities / university hospitals
8. Technology / IT companies
9. Manufacturers of fitness/health products (e.g. sports equipment, fitness bracelets, etc.)
10. Research companies (e.g. Max Planck Institute, Helmholtz Society)

*Filter: If in Q34 item in row=3 (pharmaceutical company) = 1-3 (don't entrust any data)*

Q33) You indicated that you <u>would not</u> **trust pharmaceutical companies** with **your data** <u>under any circumstances</u>. Why did you submit this rating? *OQ

*Filter: If in Q34 item in line = 3 (pharmaceutical company) = 4-5 (possibly entrust data)*

Q34) You indicated that you <u>might</u> **trust pharmaceutical companies** with **your data**. Why did you submit this rating? *OQ

*Filter: If in Q34 item in row=3 (pharmaceutical company) = 6-7 (in any case entrust data)*

Q35) You stated that you would <u>definitely</u> **entrust your data** to **pharmaceutical companies**. Why did you submit this rating? *OQ

*Filter: Display only if in Q34=Item 5 ('central database') = scale value 5.6 or 7 was selected.*

Q36) You stated that you may / you would entrust your data to a **central database of the Federal Ministry of Health**. Who do you think should have access to this database?

*Tick all groups that you think should have access. *MP, RD*

1. Ministry staff
2. Health insurance companies
3. Health authorities (e.g. local health department)
4. Pharmaceutical companies
5. Digital health start-ups
6. Universities / university hospitals
7. Technology / IT companies
8. Manufacturers of fitness/health products (e.g. sports equipment, fitness bracelets, etc.)
9. Research companies (e.g. Max Planck Institute, Helmholtz Society)
10. None of the above **EX*

*Filter: Display only if in Q34=Item 3 ('pharmaceutical company') = scale value between 1-5 was selected.*

Q37) You were rather sceptical when it comes to **pharmaceutical companies** and the use of health data. What would pharmaceutical companies have to do, prove and ensure in order to gain your trust, so that it would be also conceivable for you to pass on your data? **OQ*

*Filter: Display only if in Q34=Item 3 ('pharmaceutical company') = scale value 6 or 7 was selected.*

Q38) You have given a positive rating when it comes to providing your data to a **pharmaceutical company**. What do you think **distinguishes pharmaceutical companies** so that you are willing to provide data? And what should pharmaceutical companies pay attention to in order to keep your willingness? **OQ*

Q39) To what extent do you agree with the following statement:
"If I respond to a drug from a pharmaceutical company in the event of a serious illness and it helps me, I am happy to share my anonymous treatment data with this company for further research purposes.“ **SP*

1. Totally disagree
2. 
3. 
4. 
5. 
6. 
7. Totally agree

Q40) Finally, we are interested in a self-assessment from you regarding the **internet**. How do you feel about the internet in general? **SP*

1. The internet frightens me / confuses me
2. 
3. 
4. 
5. 
6. 
7. The internet is absolutely indispensable for me

Q41) What is your self-assessment regarding **digitisation** (i.e. the change in our entire everyday life through social media, virtual communication, internet shopping, robots, search engines, etc.)? What is your basic attitude towards digitisation? **SP*

1. Digitisation depletes us / leads to fewer social contacts
2. 
3. 
4. 
5. 
6. 
7. Digitisation is an enrichment / opens up new possibilities

44b) Another question about digitisation: What is your attitude here?**SC*

1. Digitisation is dangerous - everything is getting faster and more hectic and health and psyche suffer
2. 
3. 
4. 
5. 
6. 
7. Digitisation is positive - decisions and tasks can be completed faster and pressure is taken off

### Final sociodemographics

Finally, we would like to ask you a few questions about yourself.

Q42) How many people, including yourself, currently live in your household? **SP*

1. 1 person
2. 2 persons
3. 3 persons
4. 4 persons
5. 5 persons and more

Q43) Please tell us your marital status. *SP*

1. Unmarried – with partner
2. Single
3. Married
4. Divorced or widowed – with partner
5. Divorced or widowed – without partner

Q44) How many children of your own do you have? *SP*

1. 1 child
2. 2 children
3. 3 children
4. 4 children
5. 5 and more children
6. No children

Q45) How big is the place where you live? *SP*

1. Fewer than 5,000 inhabitants
2. 5,000 to fewer than 20,000 inhabitants
3. 20,000 to fewer than 100,000 inhabitants
4. 100,000 to fewer than 500,000 inhabitants
5. 500,000 inhabitants and more

Q46) How would you describe your residential area? *SP*

1. City centre
2. Urban fringe
3. Suburbs (up to 15 km from the city border)
4. Countryside

END
