## Supplement 2_Questionnaire AI in medicine for "Perception and knowledge of artificial intelligence in healthcare, therapy and diagnostics: A population-representative survey"

**Questionnaire – AI in Medicine 2021**

**Method:** Survey via online access panel
**Sample size:** n=1,000
**Country:** Germany
**Target group:** Adults
**Quota:** Nationally representative quotas for age, gender, and federal state, education (soft quota)
**Interview duration:** Ø 10 min

**Programming notes**

| | |
|---|---|
| **SP:* | *single pick* |
| **MP:* | *multiple pick* |
| **RD:* | *randomization* |
| **MA:* | *matrix* |
| **OQ:* | *open question* |
| **NQ:* | *numeric question* |
| **SC:* | *scale* |
| **RA:* | *ranking* |
| **DD:* | *drop down* |
| **EX:* | *exclusive* |
| *[SCREENOUT]* | *Not included in target group* |
| *[END OF INTERVIEW]* | *End of interview* |

**Welcome page**

Dear participants,

We are very pleased that you are participating in this survey on the subject of “Artificial Intelligence“.

The survey will take approximately 10 minutes of your time. Participation is of course voluntary and your data will be evaluated anonymously, so that no conclusions can be drawn about your person. Data will be used for market research purposes only.

If you would like to participate in the survey, please click "Next" to start the survey.

### 1. Sociodemographic & Screening

Q1) Please provide your gender. **SP*

1. Female
2. Male
3. Diverse / not specified

Q2) How old are you? **NQ*

_______ years *[Screenout <18]*

Q3) In which federal state do you live? **SP*

1. Baden-Württemberg
2. Bayern
3. Berlin
4. Brandenburg
5. Bremen
6. Hamburg
7. Hessen
8. Mecklenburg-Vorpommern
9. Niedersachsen
10. Nordrhein-Westfalen
11. Rheinland-Pfalz
12. Saarland
13. Sachsen
14. Sachsen-Anhalt
15. Schleswig-Holstein
16. Thüringen

Q4) What school qualifications do you have? **SP*

1. No certificate (or not yet)
2. Lower secondary/elementary school leaving certificate
3. Secondary school leaving certificate
4. A-levels, (technical) university entrance qualification without studies
5. Graduated from university

### 2. Introduction: Artificial Intelligence

Q5) Nowadays, the **term Artificial Intelligence** appears in a wide variety of places: in media, at work and even in the leisure sector. What do you mean by artificial intelligence? Where can Artificial Intelligence be used and what can it cause? Please state briefly what you know about AI. **OQ*

Q6) And against this background: How do you rate the topic of artificial intelligence?

From your point of view, is Artificial intelligence **something positive** that you endorse, **something neutral** that you observe with caution, or is it **something negative** that you oppose?
*Please answer using the scale below. *SC*

-3. Strong objection
-2.
-1.
0. Neutral
1.
2.
3. Strong endorsement

### 3. Artificial intelligence in general

**Artificial intelligence** is mentioned in media in a wide variety of contexts. Thus, you hear a wide variety of statements from different sides. We have compiled a few of these statements. For each of these statements, we would like to know to what extent you agree or disagree.

Q7) Artificial intelligence will soon destroy many jobs because simple work activities can soon be performed entirely by robots . **SC*

1. Totally disagree
2.
3.
4.
5.
6.
7. Totally agree

Q8) You have to be very cautious with artificial intelligence. Otherwise machines will soon decide instead of people whether you can get a loan or what insurance rate you have to pay. **SC*

1. Totally disagree
2.
3.
4.
5.
6.
7. Totally agree

Q9) Artificial intelligence makes our work more interesting: the routine is done automatically and we as humans take care of what requires real thinking. *SC

1. Totally disagree
2. 
3. 
4. 
5. 
6. 
7. Totally agree

Q10) Artificial intelligence helps to make decisions based on the evaluation of a large amount of data, the relationship of which is not recognizable or comprehensible to a human being. *SC

1. Totally disagree
2. 
3. 
4. 
5. 
6. 
7. Totally agree

Q11) Artificial intelligence strengthens companies in international competition because it leads to more productivity because routine tasks can be completed significantly faster than a human could. *SC

1. Totally disagree
2. 
3. 
4. 
5. 
6. 
7. Totally agree

Q12) Artificial intelligence is the only way to uncover very complex relationships between a wide variety of processes that overwhelm simple human understanding. And this is the only way we can make progress in technology and medicine. *SC

1. Totally disagree
2. 
3. 
4. 

5.
6.
7. Totally agree

Q13) Artificial intelligence is only as good as the people programming the underlying algorithms. That makes us dependent on a few specialists who intervene in our lives with their programming. *SC

1. Totally disagree
2.
3.
4.
5.
6.
7. Totally agree

Q14) Now we would like to ask you how you feel about the topic of artificial intelligence. Below please find contrasting pairs of attributes on a scale from -3 on the left through 0 to +3 on the right. Please tell us how you feel using the scale for each pair of attributes. A 0 means that you cannot assign yourself to one or the other attribute. The further you place yourself in the direction of -3 (left) or direction +3 (right), the more the assigned attribute would play a role for you. *SC

| - 3 | - 2 | - 1 | 0 | + 1 | + 2 | + 3 |
|---|---|---|---|---|---|---|
| Insecure | | | | | | Confident |
| Disapproving | | | | | | Curious |
| Anxious | | | | | | Calm |
| Pessimistic | | | | | | Optimistic |
| Confused | | | | | | Clearly sorted |
| Left alone | | | | | | Supported / supervised |
| Suspicious | | | | | | Trusting |
| Disinterested | | | | | | Interested |
| Disillusioned | | | | | | Inspired |

### 4. Artificial intelligence in medicine / medical research

So far we have talked about artificial intelligence in general, without diving deeply into specific areas of application. The remaining questions are now dedicated to a very specific area of application: the use of AI in medicine or medical research. When talking about artificial intelligence in the further survey questions, the content is related to the context of medicine (early detection, diagnosis, treatment of diseases) or medical research (discovery of new therapies, research on active molecules).

Q15) Have you ever heard about artificial intelligence in the **context of medicine** and/or **medical research**? **SP*

1. Yes
2. No

*Filter: All respondents who heard about AI in the context of medicine / medical research (Q15=1)*

Q16) What do you remember about artificial intelligence in the context of **medicine/medical research**? What areas of application do you have in mind? **OQ*

Q17) Below we have compiled various areas of application of artificial intelligence in the context of medicine / medical research. Which of these do you already know? **MA, Row RD*

*Columns*

1. It it known to me
2. I don‘t know

*Lines*

1. Suggestion of an optimal therapy for seriously ill people by AI to the treating physician, based on a large amounts of treatment data (big data) from other patients with the same or similar diseases.
2. Use in drug research: Finding suitable active ingredient molecules in huge databases, e.g. to be better able to fight new viruses or cancer cells.
3. Use in imaging procedures such as X-rays / CT scans to diagnose diseases: AI compares an X-ray images / CT scans with huge databases of other images and their existing diagnoses, thus enabling a reliable diagnosis even in difficult cases.
4. The patient controls the desired movement of their prosthesis with AI support: The basis is the AI-supported translation of muscle contractions into electronic signals that cause corresponding movements of the artificial body part.
5. Apps in which the patient enters symptoms, which the app then places in the context of disease symptoms from a wide variety of illnesses and provides information about the possible illness as a kind of initial diagnosis. The patient can then use this to visit the doctor.
6. Use for the early detection of a disease: AI uses laboratory values, X-ray images / CT and MRI images, findings from tissue samples and other treatment data of a patient (e.g. on medication), compares findings with databases and recognizes patterns that may lead to the early detection of a disease.

7. <u>Use of long-term data for early warning of a worsening of an illness</u>: Data on a patient's blood values, heart rhythm, mobility, speech ability, etc. are viewed over a long period of time, trends are recorded and compared with databases. The result of the simultaneous consideration of all trends leads to early disease prognosis.

*Filter: Show for all applications in Q17 (items in line) where in column=1(is known) was elected*

Q18) Please indicate how familiar you are with this/these application area(s) using the following scale: **MA, SC, Row RD*

*Columns*

1. I only know the application from hearsay
2. 
3. 
4. I have an intermediate level of knowledge
5. 
6. 
7. I have detailed knowledge

*Lines: Applications similar to Q17, if known*

Q19) How interested are you in **learning more** about AI in the context of medicine and medical research? **SC*

1. Not interested at all
2. 
3. 
4. 
5. 
6. 
7. Very interested

*Filter: All respondents who have a medium to high level of interest in the topic of AI (Q19=4-7)*

Q20) There are various ways to find information about a topic or to obtain information – also on the topic of AI in medicine / medical research. Please rate the following options according to your preference.
*Please rate the options on a scale from "1= very bad" to "7= very good".* **MA, Row RD*

*Columns*

1. Very bad
2. 
3. 
4. 
5. 
6. 
7. Very good

*Lines*

1. **Illustrated articles** with examples in news magazines (e.g. Spiegel, Stern, Focus)
2. **Detailed articles with pictures and interviews** in nature and science **journals** (e.g. Geo, PM, Bild der Wissenschaft, Spektrum der Wissenschaft)
3. **Scientific articles in specialist journals** (e.g. Progress in Artificial Intelligence; KI – Künstliche Intelligenz)
4. **Documentaries on television** as part of a relevant series (e.g. ZDF Wissen, Quarks, Terra X - Lesch & Co)
5. **Discussion as part of talk shows** with politicians, scientists, doctors and IT specialists (e.g. Bundesverband Medizininformatik)
6. **Books** like "Zukunftsmedizin" (Spiegel-Buch), Deep Medicine, Smart Hospital etc.
7. **Initiatives by the federal government** (such as "Wissenschaftsjahr" 2019 zur KI) as a website and brochure displays (e.g. in transregional trains)
8. **Online training courses** such as on ki-campus.org or udemy.com
9. **Web blogs** such as scienceblogs.de, RiffReporter.de, perspective-daily.de
10. **Audio podcasts** e.g. Dr. med. KI, KI in der Industrie (Fraunhofer), Hardware + KI (digitalkompakt), TechBriefing (Pioneer), Künstliche Intelligenz (Buxmann/Schmidt), KI-Board (Andreas Klug), KI Kompakt
11. **Video platforms** such as youtube / vimeo with formats by Mai Thi Nguyen-Kim, TED Talks (ted.com), Hallo! Zukunftsmedizin, Mensch Zukunft – Zukunft Mensch
12. **Websites of associations** such as Gesellschaft für Informatik, KI-Bundesverband
13. **Joint events of interest groups**, professional organisations, associations, industry, media such as forum-zukunftsmedizin.de

Q21) To what extent do you agree with the following statements? **MA*

*Columns*

1. Totally disagree
2. 
3. 
4. 
5. 
6. 
7. Totally agree

*Lines*

1. The application of artificial intelligence in medicine can be a real blessing because it helps to detect serious illnesses in a timely manner or to make them treatable in the first place.

2. Data protection should be revised for the use of artificial intelligence in medicine: no general rejection of the sharing and use of health data, but strict monitoring that prevents misuse but enables work with the data.
3. More information should be provided about the possible applications of artificial intelligence in medicine, because it offers a great opportunity for improved therapies for all of us. However, far too little is known about this.
4. If my anonymised health data helped set up databases that artificial intelligence can then use to improve diagnoses and therapies for the seriously ill, I would be willing to share my own data.

### 5. Final sociodemographics

Finally, we would like to ask you a few questions about yourself.

Q22) How many people, including yourself, currently live in your household? **SP*

1. 1 person
2. 2 persons
3. 3 persons
4. 4 persons
5. 5 persons and more

Q23) Please tell us your marital status. **SP*

1. Unmarried – with partner
2. Single
3. Married
4. Divorced or widowed – with partner
5. Divorced or widowed – without partner

Q24) How many children of your own do you have? **SP*

1. 1 child
2. 2 children
3. 3 children
4. 4 children
5. 5 and more children
6. No children

Q25) How big is the place where you live? **SP*

1. Fewer than 5,000 inhabitants
2. 5,000 to fewer than 20,000 inhabitants
3. 20,000 to fewer than 100,000 inhabitants
4. 100,000 to fewer than 500,000 inhabitants
5. 500,000 inhabitants and more

Q26) How would you describe your residential area? **SP*

1. City centre
2. Urban fringe
3. Suburbs (up to 15 km from the city border)
4. Countryside

END
